# A spatial model of CoVID-19 transmission in England and Wales: early spread and peak timing

**DOI:** 10.1101/2020.02.12.20022566

**Authors:** Leon Danon, Ellen Brooks-Pollock, Mick Bailey, Matt Keeling

## Abstract

**Background:** An outbreak of a novel coronavirus, named CoVID-19, was first reported in China on 31 December 2019. As of 9 February 2020, cases have been reported in 25 countries, including probable cases of human-to-human transmission in England.

**Methods:** We adapted an existing national-scale metapopulation model to capture the spread of CoVID-19 in England and Wales. We used 2011 census data to capture population sizes and population movement, together with parameter estimates from the current outbreak in China.

**Results:** We predict that a CoVID-19 outbreak will peak 126 to 147 days (∼4 months) after the start of person-to-person transmission in England and Wales in the absence of controls, assuming biological parameters remain unchanged. Therefore, if person-to-person transmission persists from February, we predict the epidemic peak would occur in June. The starting location has minimal impact on peak timing, and model stochasticity varies peak timing by 10 days. Incorporating realistic parameter uncertainty leads to estimates of peak time ranging from 78 days to 241 days after person-to-person transmission has been established. Seasonal changes in transmission rate substantially impact the timing and size of the epidemic peak, as well as the total attack rate.

**Discussion:** We provide initial estimates of the potential course of CoVID-19 in England and Wales in the absence of control measures. These results can be refined with improved estimates of epidemiological parameters, and permit investigation of control measures and cost effectiveness analyses. Seasonal changes in transmission rate could shift the timing of the peak into winter months, which will have important implications for health-care capacity planning.

## Introduction

An outbreak of a novel coronavirus, recently renamed CoVID-19, was first reported from Wuhan, China on 31 December 2019. During January 2020, the outbreak spread to multiple cities in China, and the first cases started appearing outside China. By the end of January 2020, 9,720 cases had been confirmed in China, with 106 confirmed cases outside China across 19 different countries(1).

Epidemiological analysis of the outbreak was quickly used to start estimating epidemiologically-relevant parameters, such as the basic reproduction number, the serial interval, the incubation period and the case fatality rate(2–7). Initial estimates suggested that the reproduction number was between 2 and 3 and the case fatality rate was less than 4%(8). Control of spread by contact tracing and isolation appears to be challenging, given what is currently known about the virus (9).

Mathematical models are useful tools for understanding and predicting the possible course of an outbreak, given a set of underlying assumptions. Here, we adapt a metapopulation model of disease transmission in England and Wales to capture the spread of CoVID-19(10). The aim is to provide predictions about the likely timing of the peak of the epidemic in England and Wales and spatial features of spread.

## Methods

### Model description

We use an existing national-scale stochastic metapopulation model of disease transmission in England and Wales. The model structure is based on the metapopulation model described in detail in Danon et al (2009)(10). In this model, the population is divided into electoral wards. Because of the changes in data availability, we restricted the model to England and Wales, whereas the original model covered Great Britain.

### Movement between wards

Transmission between wards occurs via the daily movement of individuals. For each ward, we assume that individuals contribute to the force of infection in their “home” ward during the night and their “work” ward during the day. See Danon et al 2009 for further details(10).

### Population and movement data

Data for population and movement of individuals come from the 2011 census of the United Kingdom. The population size of each of the 8,570 electoral wards is available directly from the Office of National Statistics (ONS) website. The number of individuals moving between locations is also available from the ONS website, but at the level of census output areas (OAs). We aggregated the data from OA level to electoral wards level. Spatial location of electoral ward centres are extracted from maps available from the ONS websites.

### CoVID-19 specific parameters

We use a Susceptible-Exposed-Infectious-Infectious-Recovered (SEIIR) model within each ward to capture the progression of disease within an individual (figure 1). Initial analyses used SARS-like parameters for the incubation period and infectious period, which now appear to differ from CoVID-19(4). Li et al (2020) analysed data on 425 cases reported in Wuhan in China and fitted a log-normal distribution to the incubation period, and a gamma distribution for the serial interval(2). The infectious period for SARS was estimated as the serial interval minus the incubation period, but as Li et al did not report the correlation between incubation period and serial interval, we were not able to estimate the infectious period distribution from the data, but used a uniform distribution between 2 and 3 days, to give a mean serial interval of approximately 7-8, in line with current estimates. We used two infectious states to represent a mildly symptomatic or prodromal period and a period with more pronounced symptoms. In the absence of data on the relative magnitude of these two infections states, we assumed the same length of time in each infectious state and assumed that each state was equally infectious. We sampled from each of the distributions 100 times independently (Table 1).

**Table 1:** Biological parameters and distributions used in the model.

| Parameter | Values and distribution | Reference |
| --- | --- | --- |
| Incubation period | lognorm(meanlog=log(5.2),sdlog=0.35) | Li et al(2) |
| Reproduction number | gamma(scale=2.2/100,shape=100) | Li et al(2) |
| Infectious period | uniform(2, 3) | Estimated from the mean serial interval (7.5 days) minus the mean incubation period (5.2 days) from Li et al(2) |

**Figure 1:**
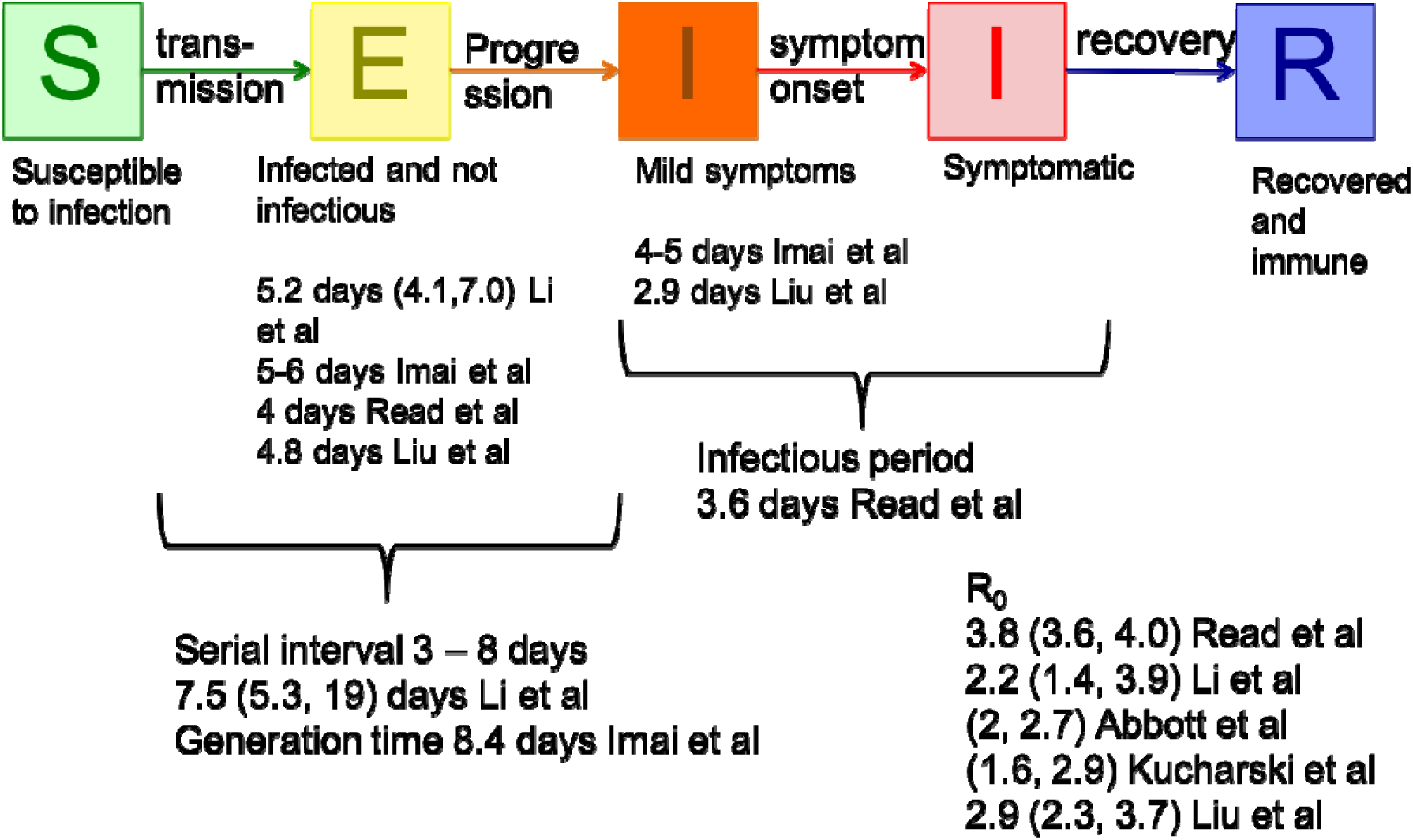
Model structure within each ward, together with associated parameters estimated from the literature.

### Initialisation and baseline model

The census data are used to initialise the population sizes within each of the 8,570 wards. At the start of the model, all individuals are assumed to be susceptible to infection with no underlying immunity in the population. We investigated a range of starting scenarios by seeding the infection in example wards London, Birmingham, Brighton, Sheffield and Cardiff. To seed infection in a ward, we move five individuals (non-commuters) from the susceptible compartment to the first infectious state.

### Impact of seasonality

We investigated the impact of a seasonally affected transmission rate, to capture potential decreased transmission during the summer months. We captured seasonal transmission by replacing the constant transmission rate with a time-varying transmission rate given by:

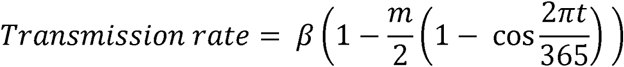

Where *m* is the magnitude of the seasonal difference in transmission, ranging from *m* = 0 (no seasonality) to *m* = 1 (maximum seasonality with no transmission at the peak of the summer).

### Epidemic characteristics

From the model, we extracted the total number of infections per day, as the number of individuals in both of the Infectious states, and the number of infected wards per day as the total number of wards with at least one individual in one of the two Infectious states. The spatial growth of the epidemic in England and Wales was visualised using interactive maps. We estimated the timing of the epidemic peak from the aggregated epidemic curve and calculated 95% prediction intervals from the model simulations.

### Implementation and data availability

The model is coded in C and is available on github (http://github.com/ldanon/MetaWards). The data are freely available from the Office for National Statistics website, or can be downloaded with the code at the github repository.

## Results

We predict that, in the absence of any interventions, a disease with “best-guess” CoVID-19-like parameters will peak a median of 133 days (range 126 - 147 days) following the start of person-to-person transmission in England and Wales. Intrinsic model stochasticity is responsible for variation between model runs. Using exactly the same parameters and seeding the infection in the same initial wards resulted in a difference in peak timing of +/- 10 days (figure 2). The attack rate for best-guess parameters had a median of 45799874 (81.67% range 81.64-81.69), with a peak incidence median 1,116,692.

**Figure 2:**
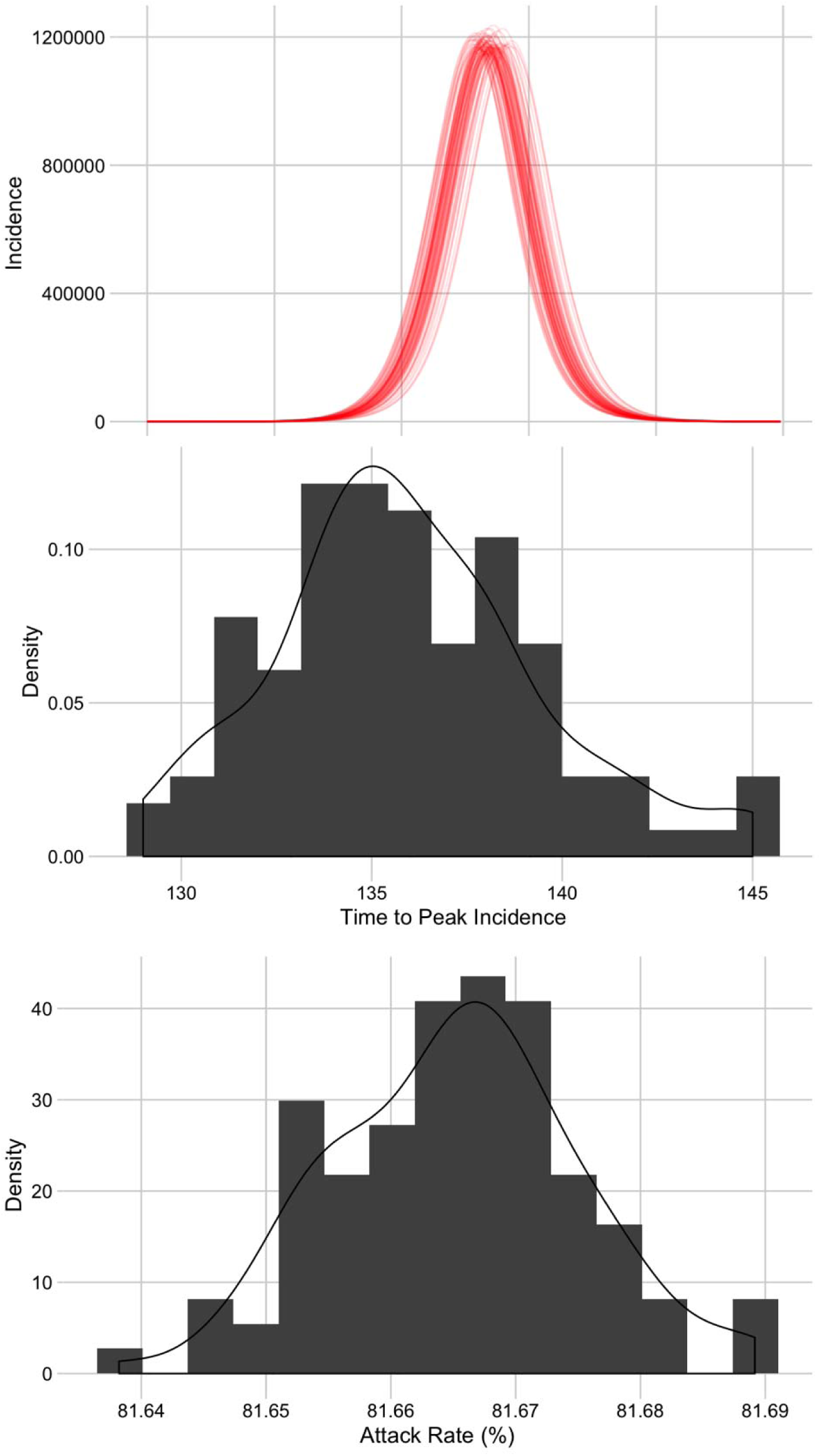
The number of cases of CoVID-19 in England and Wales in the absence of any control measures, 100 realisations of the spatial model, seeded in Brighton, using best-guess parameters from Li et. al. (top) Daily infection dynamics. (middle) The distribution of predicted time to peak incidence. (bottom) The distribution of predicted attack rate.

The initial location of cases had some, but limited impact on the timing of the epidemic in England and Wales. Epidemics seeded in Brighton, London, Birmingham and Sheffield resulted in synchronised epidemics in England, reaching urban areas first followed by rural areas. Epidemics started in Cardiff had a slower time to peak but still resulted in a generalised outbreak.

Spatially, some disaggregation between England and Wales regions is observed. An outbreak starting in Brighton, (South East England) peaks in London and the South East first, and North East England, Yorkshire and Humber and Wales last, with a ten-day lag between regions (Figure 3).

**Figure 3:**
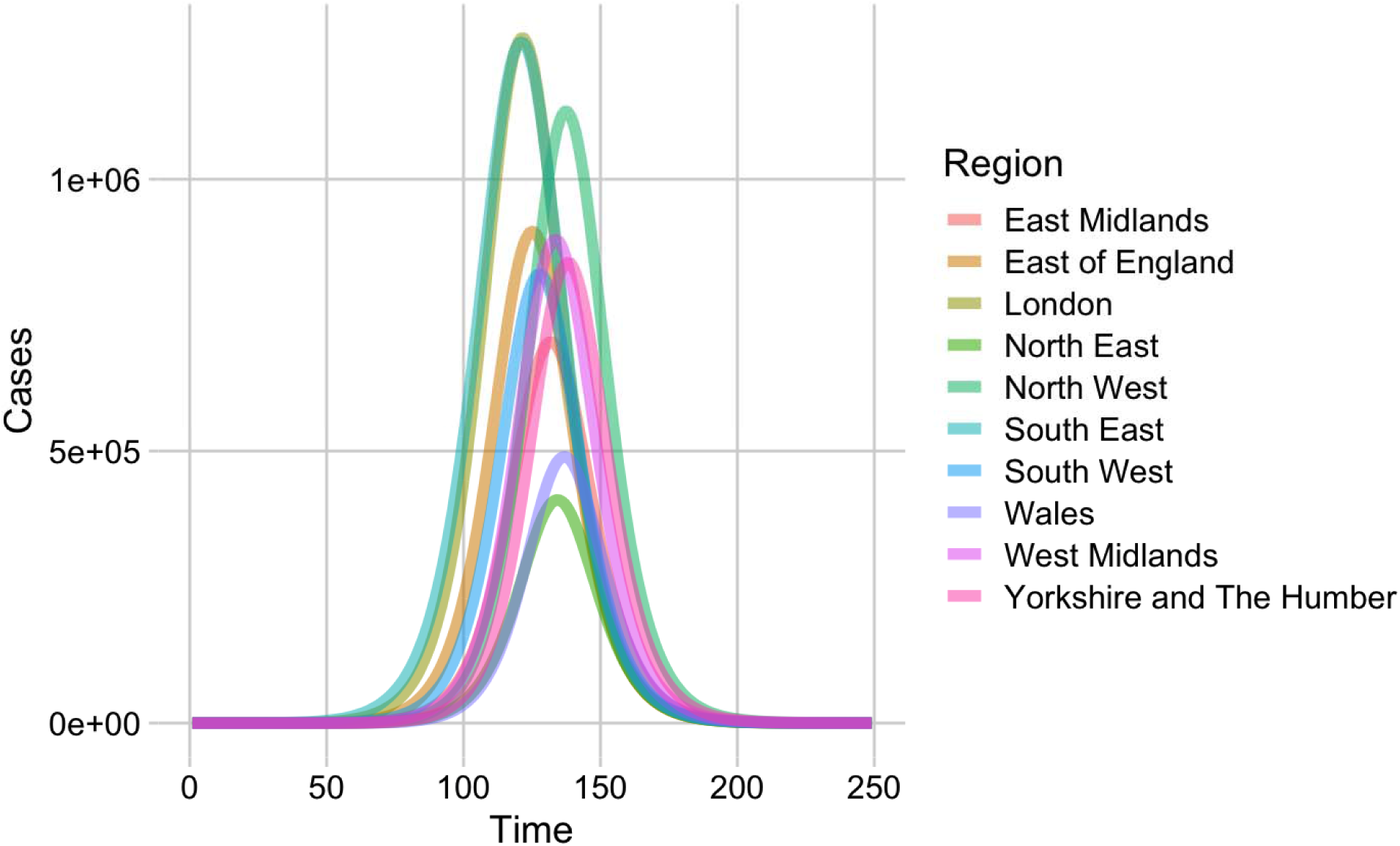
Predicted epidemic curves for a CoVID-19 outbreak broken down by region for England and Wales.

Model predictions are highly sensitive to parameter values and incorporating parameter uncertainty increases the variability of model predictions. In the absence of any control measures, all predictions resulted in epidemics that peaked within a year from the start of person-to-person transmission in England and Wales. Estimates of peak time ranged from 78 days to 241 days (Figure 4).

**Figure 4:**
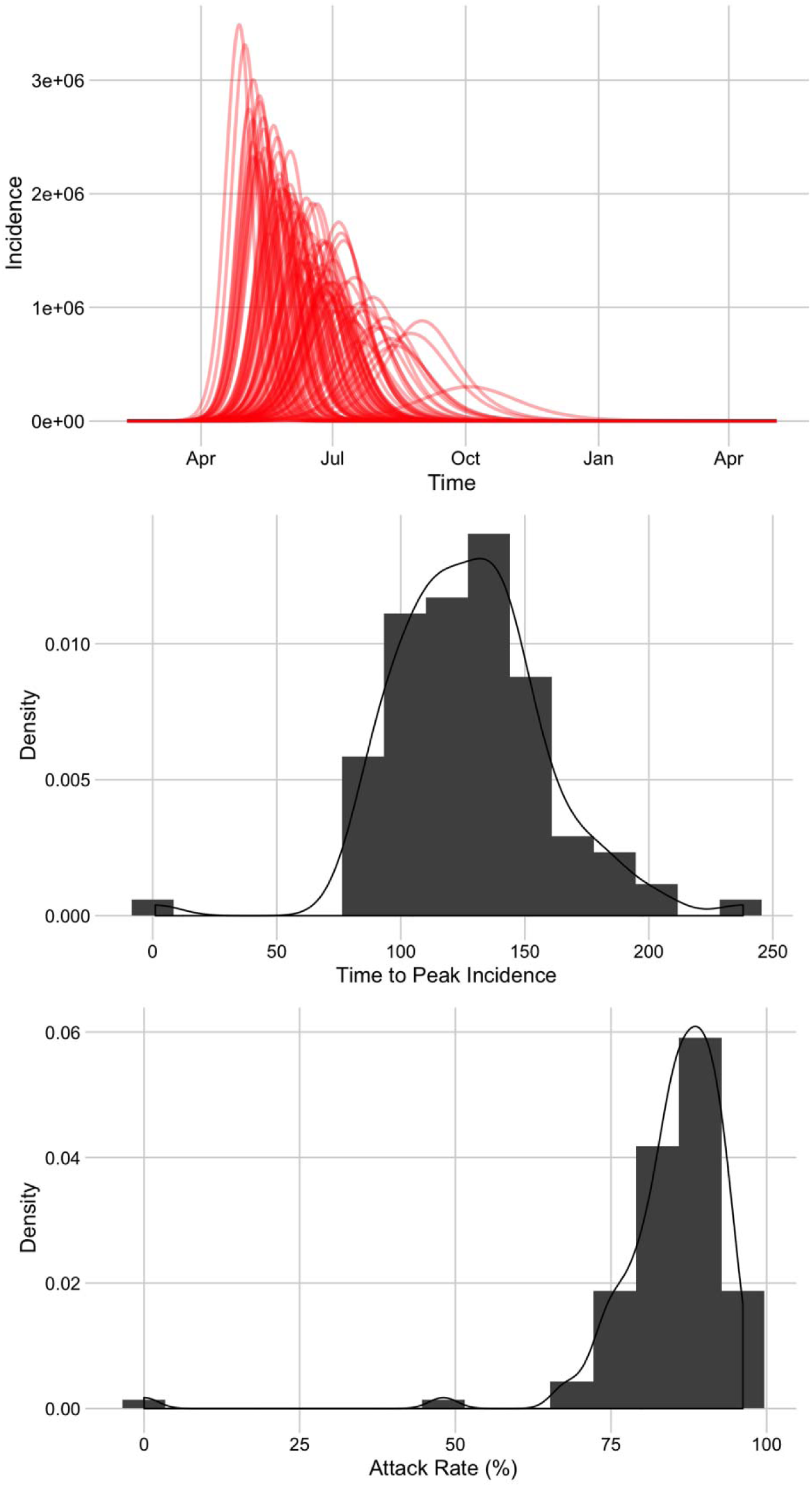
The variability in predicted epidemic curves for a Covid-19 outbreak in England and Wales, seeded in Brighton, in the absence of any control measures incorporating measured parameter uncertainty. (top) Daily infection dynamics. (middle) The distribution of predicted time to peak incidence. (bottom) The distribution of predicted attack rate.

However, seasonality in transmission has a large impact on epidemic timing, peak incidence, and final attack rates. Assuming no difference in transmission rate during the year leads to a single large epidemic after approximately 4 months (June time if transmission starts in February), as above. With a 25% reduction in transmission the epidemic is smaller and peaks later, reducing the overall attack rate by 20%. A 50% reduction in transmission results in a smaller epidemic before the summer, followed by a resurgence in cases in the following winter. The attack rate is 10% less than a non-seasonal epidemic. A 75% reduction in transmission over the summer resulted in a delayed large outbreak, but with a similar attack rate. If transmission decreases to zero over the summer then the resulting outbreak is much reduced, with an attack rate of less than 1% (Figure 5, Table 2).

**Table 2:** Effect of seasonal variation on the timing (shown in days following initial seeding), the height of the peak, and the final attack rate.

| Seasonal Term | Timing of Peak | Incidence at Peak | Attack Rate |
| --- | --- | --- | --- |
| 0 | 139 | 1172819 | 81.9 |
| 0.25 | 159 | 615599 | 65.0 |
| 0.50 | 343 | 330311 | 69.4 |
| 0.75 | 375 | 1227280 | 80.3 |
| 1.0 | 100 | 6547 | 0.53 |

**Figure 5:**
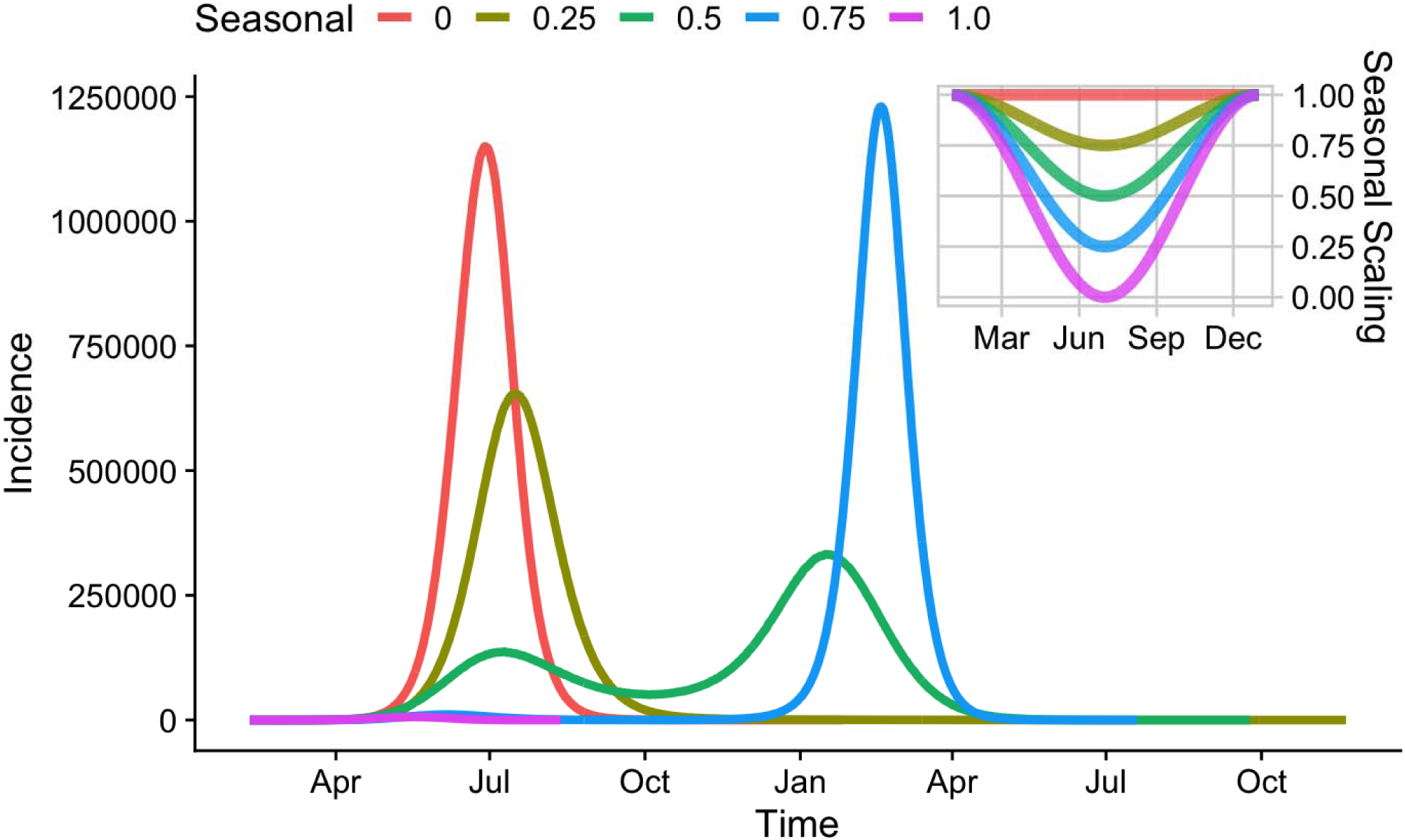
Effect of seasonal changes in transmission rate, assuming a reduction in transmission over the summer. (main panel) Incidence over time, for different values of seasonal scaling. (inset) Variation of scaling term for the course of one year, with transmission being at its lowest in July.

## Discussion

We predict that, in the absence of control measures and with no seasonality in transmission, the introduction of CoVID-19 in England and Wales has the potential to result in a synchronised outbreak that peaks at around 4 months following the start of person-to-person transmission. Our findings suggest that the height of the epidemic and the attack rate is highly dependent on seasonality of transmission and that even small changes in transmission risk can lead to large changes in attack rate due to the spatial disaggregation of the population at risk.

A combination of control measures and seasonal changes in transmission rate could shift the peak of the outbreak to the winter of 2020/21, with little effect on the final attack rate. If contact tracing and isolation efforts succeed in reducing transmission, but are unable to control the epidemic (9), an additional influx of severe CoVID-19 cases may exacerbate existing challenges with winter healthcare demand. A careful analysis of the impact of control measures on the timing of incidence of severe cases is warranted.

The strength of this model lies in the spatial heterogeneity which tempers transmission. As a comparison, an equivalent non-spatial model results in the epidemic peaking after 34 days, nearly four times faster than this spatial model, and would be unable to capture the interaction between spatial transmission and seasonality. The estimated total number of people infected in the spatial model is marginally smaller than for a non-spatial model, as the infection has the opportunity to die-out in local parts of the country. As the model framework was developed and published in 2009, it was possible to re-deploy the model for these new circumstances; developing such a model from scratch during an outbreak would be a significant challenge.

A key element missing from our model is morbidity, mortality and the treatment of cases. The model in its current form predicts the total number of infections in the community rather than diagnosed cases. Observations from China suggest that many cases have mild symptoms and that only around 5% of cases have been reported and diagnosed (3). The parameter estimates we used from China appear to be substantially different to previous coronaviruses (6). Should CoVID-19 continue spreading the UK it will become possible to get UK-specific parameter estimates and improve prediction accuracy.

As with all modelling, it is impossible to capture the full complexity of an epidemic. In this model, the major assumptions are that we have assumed that there is no change in behaviour during the course of the epidemic. In practice, as the epidemic starts spreading in England and Wales there may well be a systematic change in behaviour as was seen during the H1N1 influenza pandemic in 2009. We have not included any age-effects, such as differential mixing, susceptibility or infectiousness. That means that we are not able to investigate the impact of school closures or the impact of the summer holidays, which had a large impact on the H1N1 influenza pandemic in 2009.

## Data Availability

All the data is publicly available through the Office of National Statistics, UK, and published/preprint papers.

http://github.com/ldanon/MetaWards

## Acknowledgements

EBP was funded by the National Institute for Health Research Health Protection Research Unit (NIHR HPRU) in Evaluation of Interventions at University of Bristol in partnership with Public Health England (PHE). The views expressed are those of the author(s) and not necessarily those of the NHS, the NIHR, the Department of Health or Public Health England. LD gratefully acknowledges the financial support of The Alan Turing Institute under the EPSRC grant EP/N510129/1.

